# Increased Oxidation Susceptibility of HDL Particles as a Mechanistic Signature of Major Depressive Disorder

**DOI:** 10.64898/2025.12.02.25341460

**Authors:** Abbas F. Almulla, Mengqi Niu, Yueyang Luo, Tangcong Chen, Chenkai Yangyang, Yingqian Zhang, Michael Maes

**Author notes:** Corresponding Authors: Prof. Dr. Michael Maes and Yingqian Zhang. Sichuan Provincial Center for Mental Health Sichuan Provincial People’s Hospital, School of Medicine, University of Electronic Science and Technology of China, Chengdu 610072, China. Co-joint first author (Abbas F. Almulla, Mengqi Niu). Michael Maes https://scholar.google.co.th/citations?user=1wzMZ7UAAAAJ&hl=th&oi=ao Highly cited author: 2003-2023 (ISI, Clarivate) Scholar GPS: Worldwide #1 in molecular neuroscience; #1/4 in pathophysiology Expert worldwide medical expertise ranking, Expertscape (December 2022), worldwide: #1 in CFS, #1 in oxidative stress, #1 in encephalomyelitis, #1 in nitrosative stress, #1 in nitrosation, #1 in tryptophan, #1 in aromatic amino acids, #1 in stress (physiological), #1 in neuroimmune; #2 in bacterial translocation; 3 in inflammation, #4-5: in depression, fatigue, and psychiatry.

## Abstract

**Background:** Major depressive disorder (MDD) involves disturbances in neuroimmune–metabolic and oxidative stress (NIMETOX) pathways. However, oxidized HDL (OxHDL) and oxidized LDL (OxLDL) have not been examined together.

**Methods:** Serum OxHDL, OxLDL, and a panel of oxidative, antioxidant, and acute phase inflammatory (API) biomarkers were measured in 125 Chinese MDD patients and 40 healthy controls using ELISA and spectrophotometry.

**Results:** MDD patients showed increased OxHDL, reduced OxLDL, and markedly lowered antioxidant defenses, while classical lipid peroxidation markers remained unchanged. These alterations were independent of metabolic syndrome. The acute phase response was closely linked to reductions in HDL-related antioxidants and OxLDL. A combined biomarker model including the HDL/OxHDL ratio, apolipoprotein (ApoA)1, OxHDL, OxLDL, lipid hydroperoxides, and API index achieved an area under the ROC curve of 0.915 (SE=0.023) and a cross-validated sensitivity of 83.1% with 84.6% specificity. The variance in overall severity of depression, physiosomatic symptoms and recurrence of illness was to a large extent explained by oxidative/antioxidant biomarkers. The top-most important biomarkers were OxHDL/OxLDL (increased) and antioxidant (decreased) levels.

**Conclusion:** Increased OxHDL is a key component of MDD, indicating structural and dysfunctional HDL particles and oxidative damage to its major structural protein (ApoA1). HDL particles rather than LDL particles and other lipids are the most vulnerable sites to be attacked by oxidative stress and inflammatory processes in MDD.

These data support the view that increased oxidative damage to HDL particles is a key process in MDD. Preventing HDL particle oxidation is a major new drug target in MDD.

## Introduction

Major depressive disorder (MDD) is recognized as a condition marked by heightened oxidative and nitrosative stress (ONS), indicated by increased levels of reactive oxygen and nitrogen species (RONS) alongside diminished antioxidant defenses (Maes, Galecki et al. 2011, Maes, Almulla et al. 2025). These aberrations are part of interconnected neuroimmune, metabolic and oxidative stress (NIMETOX) pathways (Maes, Galecki et al. 2011, Maes, Almulla et al. 2025). Recent findings indicate that MDD is associated with abnormal lipid metabolism and increased lipid peroxidation, evidenced by elevated levels of malondialdehyde (MDA), 4-hydroxynonenal, peroxides, 8-isoprostanes, and a higher apolipoprotein (Apo)B/ApoA1 ratio (Almulla, Thipakorn et al. 2023, Jirakran, Almulla et al. 2025). These changes coincide with impaired reverse cholesterol transport (RCT) and decreased levels of lipid-associated antioxidants, including paraoxonase-1 (PON1), lecithin–cholesterol acyltransferase (LCAT), high-density lipoprotein (HDL), and ApoA1 (Almulla, Thipakorn et al. 2023, Jirakran, Almulla et al. 2025). Moreover, elevated levels of advanced oxidation protein products (AOPPs) indicate significant chlorinative stress and oxidative damage to proteins (Carpita, Betti et al. 2021).

MDD consistently demonstrates decreased serum HDL levels and diminished RCT (Almulla, Thipakorn et al. 2023, Jirakran, Almulla et al. 2025). Increased myeloperoxidase (MPO) is a significant factor in this impairment, as it oxidizes ApoA1 and facilitates the formation of dysfunctional HDL (Shao, Pennathur et al. 2012, Cooke, Morris et al. 2018). ApoA1 possesses antioxidant properties and is diminished in MDD, leading to a further weakening of the HDL-associated antioxidant system (Chen, Niu et al. 2025). Further alterations, including glycation and carbamylation, may undermine HDL integrity (Sun, Yang et al. 2016, Gomes Kjerulf, Wang et al. 2020).

In MDD, there is no data on oxidized HDL (OxHDL) or the HDL to OxHDL ratio, despite established oxidative mechanisms. The significance of this gap lies in the likelihood that oxidative damage contributes to the decreased levels of circulating HDL and ApoA1 frequently observed in MDD. Conversely, oxidized LDL (OxLDL) is documented in MDD, with multiple studies and meta-analyses indicating increased levels of OxLDL (Maes, Kubera et al. 2013, Almulla, Thipakorn et al. 2023, Solak, Akkuş et al. 2025). Oxidized LDL may generate immunogenic neoepitopes, and previous research indicated notable increases in IgG levels targeting OxLDL (Almulla, Thipakorn et al. 2023).

Both metabolic syndrome (MetS) and obesity are linked to increased oxidative stress and diminished antioxidant defenses (Maes, Almulla et al. 2025). The redox imbalance is significant in MetS, probably attributed to the synergistic effects of MetS components such as insulin resistance and dyslipidemia (Morelli, Maes et al. 2021). MetS often coexists with MDD (de Melo, Nunes et al., 2017). Immune and ONS data in MDD and comorbid MetS show that not omitting or controlling for possible effects of MetS may lead to erroneous deductions concerning NIMETOX data (de Melo, Nunes et al. 2017, Morelli, Maes et al. 2021, Jirakran, Vasupanrajit et al. 2025).

MetS and obesity are significantly less common in Chinese populations compared to Western cohorts, making biomarker studies in China valuable for understanding the fundamental pathophysiology of MDD independent of MetS or obesity (Hirode and Wong 2020, Emmerich, Fryar et al. 2024). Recent data from China indicate ongoing NIMETOX abnormalities after controlling for MetS and body mass index (BMI) or omitting MetS or obesity patients, implying that these changes represent inherent disease mechanisms rather than secondary metabolic effects (Chen, Niu et al. 2025, Luo, Niu et al. 2025).

ONS is closely associated with the acute phase response in MDD (Liu, Chen et al. 2012). Acute phase proteins (APPs) such as albumin, transferrin, and haptoglobin, serve as antioxidant buffers (Maes, Galecki et al. 2011). Almulla et al. found that monomeric C-reactive protein (mCRP), in conjunction with negative APPs such as albumin and transferrin, effectively differentiates MDD patients from control subjects with a high accuracy (Almulla, Niu et al. 2025). Furthermore, Chen et al. demonstrated that a combination of an APP-inflammatory (API) index with metabolic markers, including total cholesterol, BMI, and ApoA1, showed elevated diagnostic accuracies according to different regression models (Chen, Niu et al. 2025). The findings suggest that diagnostic performance is enhanced when ONS biomarkers are assessed in conjunction with API indices, aligning with the NIMETOX model of MDD.

Niu et al. (Niu, Zhang et al. 2025) have shown that the phenome of MDD includes integrated domains of affective, somatic, vegetative, melancholic, and suicidal symptoms. The Overall Severity of Depression (OSOD) represents the overarching (general) factor that underlies these domains, while a single group factor reflects the physiosomatic dimension encompassing specific fatigue-related and physical symptoms. Apart from these two factors, the Recurrence of Illness (ROI) should also be used to assess the clinical picture of MDD (Niu, Zhang et al. 2025). ROI quantifies the lifetime burden of illness associated with depressive episodes and suicidal behaviors (Maes, Jirakran et al. 2024). Moreover, increased adverse childhood experiences (ACEs) may affect the phenome of depression as well as NIMETOX pathways (Maes, Almulla et al. 2025). Nevertheless, no studies have examined the associations between OxHDL/OxLDL and the general phenome factor, the single-group physiosomatic factor, ROI and ACEs.

Hence, this study aims to examine oxidatively modified HDL and LDL particles in Chinese patients with MDD. Additionally, it measures HDL-related components (ApoA1, CMPAase), lipid peroxidation markers (LPO, MDA), chlorinative stress indices (MPO, AOPP), nitrosative products (NOx), total oxidative status (TOS), total antioxidant capacity (TAOC), and negative (albumin, transferrin), and positive (mCRP) acute phase proteins. Additionally, we will evaluate the effects of the AP inflammatory response and MetS on the OS and ANTIOX biomarkers.

## Material and Methods

### Participants

This study utilized a cross-sectional, case–control design at the Psychiatric Center of Sichuan Provincial People’s Hospital in Chengdu, China. A total of 165 participants were recruited, comprising 125 patients diagnosed with MDD and 40 healthy controls. Patients were enrolled consecutively from clinical services, while controls were selected from hospital staff, their relatives, and acquaintances of the patients. Controls were matched to patients based on age, sex, education, and body mass index to reduce demographic confounding.

Participants eligible for the study were aged between 18 and 65 years, regardless of sex, and had provided written informed consent before participation. The inclusion criteria for patients mandated a diagnosis of MDD as per the Diagnostic and Statistical Manual of Mental Disorders, Fifth Edition (DSM-5), along with a Hamilton Depression Rating Scale (HAMD-21) (Hamilton 1960) score exceeding 18. Exclusion criteria included the presence of comorbid psychiatric conditions such as bipolar disorder, schizophrenia, schizoaffective disorder, psycho-organic syndromes, autism spectrum disorders, eating disorders, and substance use disorders (excluding nicotine dependence). Individuals with borderline and antisocial personality or developmental disorders, including significant intellectual disability, were excluded from the study. Individuals diagnosed with neurological conditions, including stroke, epilepsy, brain tumors, Parkinson’s disease, Alzheimer’s disease, or multiple sclerosis, were excluded from eligibility. Other medical exclusions comprised autoimmune or inflammatory diseases such as systemic lupus erythematosus, inflammatory bowel disease, rheumatoid arthritis, and psoriasis, as well as type 1 diabetes mellitus, chronic obstructive pulmonary disease, and malignancies.

Additional exclusion criteria for all participants encompassed pregnancy or lactation, a history of severe allergic reactions within the past month, active or recent infections within the last three months, and treatment with immunosuppressive or immunomodulatory medications, including glucocorticoids. Individuals receiving therapeutic doses of antioxidants or omega-3 fatty acids, those who had surgery within the last three months, and those with regular analgesic use were excluded from the study. Moreover, healthy controls were excluded if they had a current or past diagnosis of MDD, dysthymia, or anxiety disorders as per DSM-5 criteria, or if there was a family history of affective disorders, suicide, schizophrenia, or substance use disorders excluding nicotine dependence.

### Clinical assessments

All participants underwent a clinician-led, semi-structured interview that captured demographic and clinical information, including age, sex, education, income, illness course, medical comorbidities, number and timing of depressive episodes, personal history, and family history. Psychiatric diagnoses were established with the Mini International Neuropsychiatric Interview (M.I.N.I.), which screens current and lifetime DSM-IV/ICD-10 disorders and includes assessment of antisocial personality disorder. Disorders covered encompassed MDD and dysthymia episodes, hypomania, suicidal ideation and behavior, panic disorder, agoraphobia, social anxiety disorder, generalized anxiety disorder, obsessive–compulsive disorder, post-traumatic stress disorder, alcohol, and non-alcohol substance use disorders, psychotic disorders, anorexia nervosa, and bulimia nervosa.

On the same visit, the same evaluator rated symptom severity. The different scales used to compute OSOD, ROI and physiosomatic symptoms and the calculation methods were described in detail by Niu et al. (2025). A ROI index was derived as the z-sum of three standardized counts: lifetime number of depressive episodes, number of suicide attempts (up to one month before the index episode), and number of suicidal ideations (up to one month before the index episode). Scores for emotional and physical abuse, sexual abuse, emotional neglect, and physical neglect were computed in accordance with previous research (Vasupanrajit, Maes et al. 2024). Furthermore, sexual abuse was included in the analyses alongside the aggregate of the four additional ACEs, referred to as sum “four ACEs.”

Anthropometric and hemodynamic measurements included waist circumference, body weight, and height. BMI was calculated as kg/m². Waist circumference was measured in the horizontal plane midway between the iliac crest and the lowest rib. An adiposity composite was constructed as zBMI + zWC. Metabolic syndrome (MetS) was defined according to the 2009 joint statement criteria, requiring at least three of the following: WC ≥ 90 cm (men) or ≥ 80 cm (women), triglycerides ≥ 150 mg/dL, HDL-cholesterol < 40 mg/dL (men) or < 50 mg/dL (women), blood pressure ≥ 130/85 mmHg or antihypertensive treatment, and fasting glucose ≥ 100 mg/dL or a diabetes diagnosis. MetS rank reflected the number of fulfilled components.

### Biomarkers Quantification

Fasting venous blood (30 mL) was obtained into serum tubes using disposable syringes in the early morning window (06:30–08:00). Samples were centrifuged at 3500 rpm, the serum phase was aliquoted into Eppendorf tubes, and specimens were preserved at −80 °C until assay. The analytes quantified, the methods and equipment are listed in ESF Table 1. Composite indices were derived as follows. Total oxidative stress was calculated as the sum of the relevant oxidative stress biomarkers, namely zLPO + zMDA + zMPO + zTOS + zNOx (labeled as “Total ONS” index). The total antioxidant defense was calculated as the sum of the relevant antioxidant biomarkers, namely zAlbumin + zHDL + zApoA1 + zCMPAase + zTAOC (labeled as “Total ANTIOX”). Consequently, we computed the Total ONS/Total ANTIOX ratio as zTotal ONS – zTotal ANTIOX (labeled as zONS - zANTIOX ratio). In addition, we computed two ratios based on OxHDL, namely HDL/OxHDL as zHDL – zOxHDL (reflecting the ratio of measured HDL levels on those of OxHDL), and OxHDL/OxLDL as zOxHDL – zOxLDL (reflecting whether HDL or LDL is the preferable site of oxidative stress attacks). The AP inflammatory (API) index was computed as z(mCRP) − z(albumin) − z(transferrin).

**Table 1.** Differences in biomarkers between major depressive disorder (MDD) and healthy controls (HC)

| **Variables** | **HC**<br>**(n = 40)** | **MDD**<br>**(n = 125)** | **F value**<br>**(df = 1/154)** | **p-value** |
| --- | --- | --- | --- | --- |
| Albumin (g/L) | 43.47 (0.45) | 41.05 (0.25) | 20.44 | <0.001 |
| Transferrin (mg/dL) | 259.0 (6.2) | 229.3 (3.5) | 17.15 | <0.001 |
| Monomeric CRP* | -0.336 (0.169) | 0.149 (0.094) | 6.13 | 0.014 |
| Acute phase-inflammatory index ( z score) | -1.483 (0.298) | 0.537 (0.167) | 34.01 | <0.001 |
| LPO (μmol/L) | 4.22 (0.38) | 3.43 (0.21) | 3.78 | 0.053 |
| MDA (μmol/L) | 3.56 (0.28) | 3.79 (0.16) | 0.23 | 0.631 |
| MPO (U/mL) | 0.922 (0.292) | 0.938 (0.164) | 0.28 | 0.601 |
| AOPPs (pg/mL) | 699.8 (23.2) | 682.6 (13.0) | 0.44 | 0.508 |
| TOS (μmol/L) | 8.89 (0.51) | 9.36 (0.29) | 2.01 | 0.159 |
| NOx (μmol/L) | 40.8 (4.8) | 36.1 (2.7) | 0.96 | 0.329 |
| Total ONS (z score) | 0.298 (0.175) | 0.300 (0.119) | 0 | 0.993 |
| HDL (mmol/L) | 1.63 (0.05) | 1.36 (0.03) | 9.29 | 0.003 |
| Apolipoprotein A1 (g/L) | 1.69 (0.03) | 1.42 (0.02) | 25.03 | <0.001 |
| CMPAase (U/mL) | 16.63 (0.60) | 15.47 (0.34) | 5.22 | 0.024 |
| Total antioxidant capacity (TAOC) (U/mL) | 9.58 (0.32) | 8.35 (0.18) | 22.91 | <0.001 |
| Total ANTIOX (z score) | 0.747 (0.161) | -0.367 (0.109) | 33.32 | <0.001 |
| Total ONS / Total ANTIOX index (z score) | -0.330 (0.167) | 0.473 (0.113) | 16.11 | <0.001 |
| Oxidized HDL (OxHDL) (ng/mL) | 9.14 (1.12) | 11.88 (0.62) | 8.92 | 0.003 |
| Oxidized LDL (OxLDL) (pg/mL) | 317.4 (18.6) | 232.8 (10.45) | 16.29 | <0.001 |
| z HDL – z OxHDL (z score) | 0.308 (0.164) | -0.500 (0.111) | 16.94 | <0.001 |
| z OxHDL – z OxLDL (z score) | -0.690 (0.175) | 0.302 (0.118) | 22.50 | <0.001 |

All data are shown as marginal estimated mean (SE); results of GLM analysis with age, sex, body mass index and metabolic syndrome as covariates. \*Adjusted for age, sex, and body mass index (residual values). All significant differences shown in this table remained significant after False Discovery Rate p correction.
LPO: Lipid peroxide, MDA: Malonaldehyde, MPO: Myeloperoxidase, AOPP: Advanced oxidation protein products, TOS: total oxidative status, Nox: Nitric oxide metabolites, CMAase: chloromethyl phenyl acetate, ONS: oxidative and nitrosative stress; HDL: High density lipoprotein cholesterol, ANTIOX: antioxidant defenses, OxHDL: oxidized high-density lipoprotein, OxLDL: oxidized low-density lipoprotein.

### Data Analysis

All analyses were conducted using IBM SPSS Statistics for Windows, version 30. Central tendency measures were presented with corresponding standard deviations or, alternatively, as estimated marginal means accompanied by standard errors. Data distributions were assessed prior to analysis and, when required, normalized through log10, square-root, rank-order, or Winsorized transformations to satisfy model assumptions. Analysis of variance was employed for continuous variables, while chi-square tests were utilized for categorical measures in comparative analyses between healthy controls and patients diagnosed with MDD. We employed general linear models incorporating age, sex, and BMI as factors to evaluate differences in APPs, ONS indicators, and other pertinent variables. The False Discovery Rate (FDR) was utilized to correct the significance for the number of tests conducted.

The effectiveness of biomarkers in distinguishing control individuals from those diagnosed with MDD was assessed through binary logistic regression models. The control group was designated as the reference category, with the dependent variable defined as MDD status. The analysis incorporated factors such as age, gender, smoking status, and BMI. For each model, we provided unstandardized regression coefficients (B), standard errors (SE), Wald χ² statistics, p-values, odds ratios with 95% confidence intervals, and Nagelkerke pseudo-R² values. The Wald statistic is calculated using the formula (B/SE)². A 10-fold cross-validation linear discriminant analysis was conducted to assess the model’s generalizability and stability. Predictive performance was assessed using overall classification accuracy, the maximum Kolmogorov-Smirnov statistic, the area under the ROC curve, the Gini index, and various test statistics.

Multiple regression analyses were conducted to examine the factors that may predict acute-phase reactants and symptom rating scores. Demographic, metabolic, and biomarker variables were evaluated as potential predictors. Data entry was conducted both manually and automatically through a sequence of steps. Significance levels of p = 0.05 and p = 0.07 were employed to determine the inclusion or exclusion of variables through an automated method utilizing SPSS’s automatic linear modeling, which incorporates built-in overfitting control. Standardized beta coefficients, degrees of freedom, R², F values, and corresponding p-values were recorded for each regression model. We employed the White and modified Breusch-Pagan tests to assess heteroskedasticity and utilized tolerance and variance inflation factors to evaluate collinearity. The significance level was established at 0.05, and all tests were conducted as two-tailed.

## Results

### Socio-demographic and clinical characteristics

ESF Table 2 summarizes participant characteristics. Age, sex, and smoking did not differ between patients with MDD and controls. Metabolic indices were comparable,including MetS prevalence, MetS rank, waist circumference, BMI, and the composite zBMI + zWC. The ROI, OSOD, physiosomatic scores and ACEs were higher in MDD than in controls.

**Table 2:**
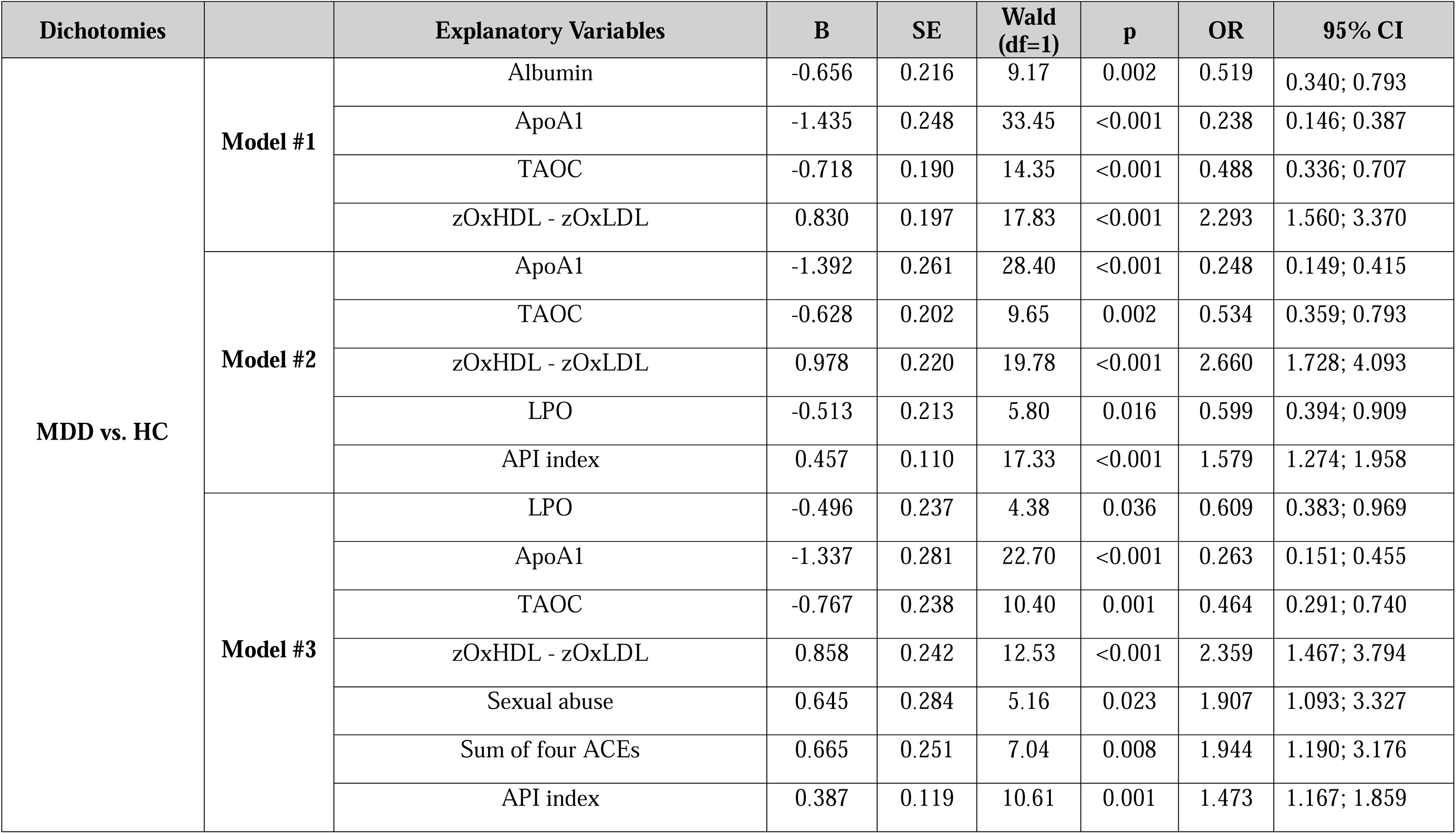
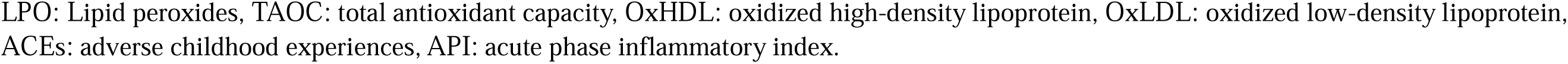
Results of binary logistic regression analyses with major depression (MDD) as dependent variable and healthy controls as reference group.

| Dichotomies |  | Explanatory Variables | B | SE | Wald (df=1) | p | OR | 95% CI |
| --- | --- | --- | --- | --- | --- | --- | --- | --- |
| MDD vs. HC | Model #1 | Albumin | -0.656 | 0.216 | 9.17 | 0.002 | 0.519 | 0.340; 0.793 |
|  |  | ApoA1 | -1.435 | 0.248 | 33.45 | <0.001 | 0.238 | 0.146; 0.387 |
|  |  | TAOC | -0.718 | 0.190 | 14.35 | <0.001 | 0.488 | 0.336; 0.707 |
|  |  | zOxHDL - zOxLDL | 0.830 | 0.197 | 17.83 | <0.001 | 2.293 | 1.560; 3.370 |
|  | Model #2 | ApoA1 | -1.392 | 0.261 | 28.40 | <0.001 | 0.248 | 0.149; 0.415 |
|  |  | TAOC | -0.628 | 0.202 | 9.65 | 0.002 | 0.534 | 0.359; 0.793 |
|  |  | zOxHDL - zOxLDL | 0.978 | 0.220 | 19.78 | <0.001 | 2.660 | 1.728; 4.093 |
|  |  | LPO | -0.513 | 0.213 | 5.80 | 0.016 | 0.599 | 0.394; 0.909 |
|  |  | API index | 0.457 | 0.110 | 17.33 | <0.001 | 1.579 | 1.274; 1.958 |
|  | Model #3 | LPO | -0.496 | 0.237 | 4.38 | 0.036 | 0.609 | 0.383; 0.969 |
|  |  | ApoA1 | -1.337 | 0.281 | 22.70 | <0.001 | 0.263 | 0.151; 0.455 |
|  |  | TAOC | -0.767 | 0.238 | 10.40 | 0.001 | 0.464 | 0.291; 0.740 |
|  |  | zOxHDL - zOxLDL | 0.858 | 0.242 | 12.53 | <0.001 | 2.359 | 1.467; 3.794 |
|  |  | Sexual abuse | 0.645 | 0.284 | 5.16 | 0.023 | 1.907 | 1.093; 3.327 |
|  |  | Sum of four ACEs | 0.665 | 0.251 | 7.04 | 0.008 | 1.944 | 1.190; 3.176 |
|  |  | API index | 0.387 | 0.119 | 10.61 | 0.001 | 1.473 | 1.167; 1.859 |

LPO: Lipid peroxides, TAOC: total antioxidant capacity, OxHDL: oxidized high-density lipoprotein, OxLDL: oxidized low-density lipoprotein, ACEs: adverse childhood experiences, API: acute phase inflammatory index.

### ONS biomarkers in MDD vs controls

**Table 1** shows biomarker differences. Albumin, transferrin, HDL, ApoA1, CMPAase, and TAOC were lower in patients than controls. The Total ANTIOX composite score and the zHDL – zOxHDL ratio were lower, together with OxLDL. By contrast, mCRP, API, OxHDL, the zONS - zANTIOX index, and the zOxHDL – zOxLDL ratio were increased. LPO, MDA, MPO, AOPP, TOS, NOx, and Total ONS did not differ from controls. These differences remained significant after FDR p-correction.

Group contrasts by MetS status are provided in ESF, Table 3. Participants with MetS showed higher TOS, higher Total ONS, and a higher zONS – zANTIOX ratio than those without MetS. HDL, ApoA1, and the zHDL – zOxHDL ratio were lower in MetS. Albumin, transferrin, mCRP, the API index, LPO, MDA, MPO, AOPP, NOx, CMPAase, OxLDL, OxHDL, Total ANTIOX and the zOxHDL – zOxLDL ratio were not significantly different between those with and without MetS.

**Table 3.** Diagnostic performance metrics of the different models shown in Table 2 in differentiating major depressed patients from healthy controls.

| Predictors | AUC ROC | Gini index | Max K-S | Overall Model Quality | Accuracy | Sens/Spec. |
| --- | --- | --- | --- | --- | --- | --- |
| Model #1. Albumin, ApoA1, TAOC, zOxHDL - zOxLDL | 0.888 (0.022) | 0.776 | 0.668 | 0.84 | 81.22% | 81.97% / 80.3% |
| Model #2. ApoA1, TAOC, zOxHDL - zOxLDL, LPO, API index | 0.906 (0.020) | 0.812 | 0.696 | 0.87 | 82.2% | 81.3% / 83.2% |
| Model #3. LOOH, ApoA1, TAOC, zOxHDL – zOxLDL, Sexual abuse, Sum of four ACEs, APP index | 0.927 (0.018) | 0.855 | 0.798 | 0.89 | 88.6% | 88.5% / 88.8% |

LPO: Lipid peroxides, AOPP: Advanced oxidation protein products, Apo: Apolipoprotein, TAOC: total antioxidant capacity, oxHDL: oxidized high-density lipoprotein, oxLDL: oxidized low-density lipoprotein, API: acute phase inflammatory index, ACEs: adverse childhood experiences.

Our findings reveal that a total of 92 patients diagnosed with MDD were treated with antidepressants. Additionally, 10 patients were given mood stabilizers, 44 received atypical antipsychotics, and 68 were prescribed benzodiazepines. The administration of these pharmacological agents did not produce any notable effects on any of the ONS and ANTIOX variables even when FDR p-value correction was not applied. The findings indicate that the interventions administered to the patients did not influence the outcome of the biomarkers in the current study. Furthermore, among the cohort diagnosed with MDD, no notable correlations were identified between the use of these medications and OSOD and physiosomatic symptoms. A notable correlation was observed between the use of mood stabilizers and ROI, characterized by a correlation coefficient of r=0.227, with a significance level of p=0.003. Nonetheless, this correlation lost its significance subsequent to the implementation of the FDR p-correction. It is reasonable to infer that this trend suggests patients demonstrating heightened ROI are frequently prescribed mood stabilizers.

### Diagnostic performance of oxidative stress biomarkers

Binary logistic regression analyses were conducted to assess the diagnostic accuracy of ONS biomarkers in distinguishing patients with MDD from healthy controls (**Table 2)**. Model #1 revealed (based on all ONS and ANTIOX biomarkers, but without the API index) that MDD was significantly associated with reduced albumin, ApoA1, and TAOC levels, and an increased zOxHDL – zOxLDL ratio (χ² = 129.895, df = 4, p < 0.001, Nagelkerke R² = 0.578). This model achieved a high overall classification accuracy as shown in **Table 3**. When the API index was added to the model (Model #2), the predictive power improved, showing that MDD is inversely associated with ApoA1, TAOC, and LPO, and positively with the zOxHDL – zOxLDL ratio and the API index. Table 3 shows that the model’s classification accuracy increased. Linear Discriminant Analysis (LDA) (automatic) showed that six discriminatory variables significantly separated MDD from controls (Wilk’s λ = 0.514, χ² = 149.854, df = 6, p < 0.001, canonical correlation = 0.697), namely ApoA1, LPO, API index, zHDL – zOxHDL, OxHDL, and OxLDL. The AUC ROC curve was 0.915 (± 0.023), GINI index = 0.830, with a cross-validated sensitivity of 83.1% and a specificity of 84.6%.

We have also entered both ACE indices and sexual abuse in the binary logistic regression analyses to ascertain that the above results are not affected by the impact of ACEs. In Model #3, the inclusion of the sum of four ACEs and sexual abuse significantly improves predictive power, indicating that the effects of ApoA1, TAOC, LPO, and API are independent from ACEs.

### Correlations between ACEs, ONS biomarkers, and API

**Table 4** displays the Pearson correlations between ACEs, the clinical phenome, psychosomatic symptoms, the API index, Total ONS, and Total ANTIOX. The sum of four ACEs exhibited notable negative correlations with the Total ANTIOX index and OxLDL. In addition, ACEs exhibited a negative correlation with ApoA1 (r = –0.261, p < 0.001) and CMPAase (r = –0.261, p < 0.001), but no correlation was found with HDL. The MDD phenome exhibited negative correlations with the Total ANTIOX index, OxLDL, and the zHDL – zOxHDL ratio, while showing positive correlations with OxHDL and the zOxHDL – zOxLDL ratio. Physiosomatic symptoms exhibited a comparable pattern, demonstrating a negative correlation with the Total ANTIOX index, OxLDL, and the zHDL – zOxHDL ratio, while showing a positive correlation with OxHDL and the zOxHDL – zOxLDL ratio. The API index exhibited a negative correlation with the Total ANTIOX index, OxLDL, and the zHDL - zOxHDL ratio, while showing a positive association with the zOxHDL - zOxLDL ratio.

**Table 4.** Intercorrelation matrix (Pearson’s correlation coefficients) between oxidative stress biomarkers, adverse childhood experiences (ACEs), the clinical phenome of depression, and the acute phase inflammatory (API) index.

| **Variables** | **Sum of four ACEs** | **OSOD** | **Physiosomatic symptoms** | **API index** |
| --- | --- | --- | --- | --- |
| **Total ANTIOX index** | -0.301 (<0.001) | -0.495 (<0.001) | -0.444 (<0.001) | -0.562 (<0.001) |
| **Total ONS index** | -0.059 (0.453) | -0.082 (0.324) | -0.094 (0.259) | -0.019 (0.812) |
| **OxHDL** | 0.042 (0.596) | 0.224 (0.006) | 0.202 (0.014) | 0.095 (0.228) |
| **OxLDL** | -0.192 (0.014) | -0.294 (<0.001) | -0.279 (<0.001) | -0.228 (0.004) |
| **zHDL - zOxHDL** | -0.130 (0.098) | -0.356 (<0.001) | -0.320 (<0.001) | -0.207 (0.008) |
| **zOxHDL - zOxLDL** | 0.154 (0.050) | 0.343 (<0.001) | 0.319 (<0.001) | 0.213 (0.007) |

OSOD: overall severity of depression; ONS: oxidative and nitrosative stress; ANTIOX: antioxidant defenses, OxHDL: oxidized high-density lipoprotein, OxLDL: oxidized low-density lipoprotein

### Effect of the API index on oxidative stress and antioxidant biomarkers

**Table 5** demonstrates that the API index exerted a broad influence on antioxidant parameters. The API index showed significant inverse associations with HDL, ApoA1, CMPAase, TAOC, Total ANTIOX score, OxLDL, and zHDL – zOxHDL, while it was positively related to the zOxHDL – zOxLDL ratio. After adjusting for the API index, MDD diagnosis remained significantly associated with HDL, ApoA1, TAOC, Total ANTIOX index, OxLDL, OxHDL, zHDL – zOxHDL, and zOxHDL – zOxLDL ratios. The effect on CMPAase was no longer significant after adjustment, whereas the association of OxHDL with MDD persisted, suggesting that changes in OxHDL are independent of the API response.

**Table 5.**
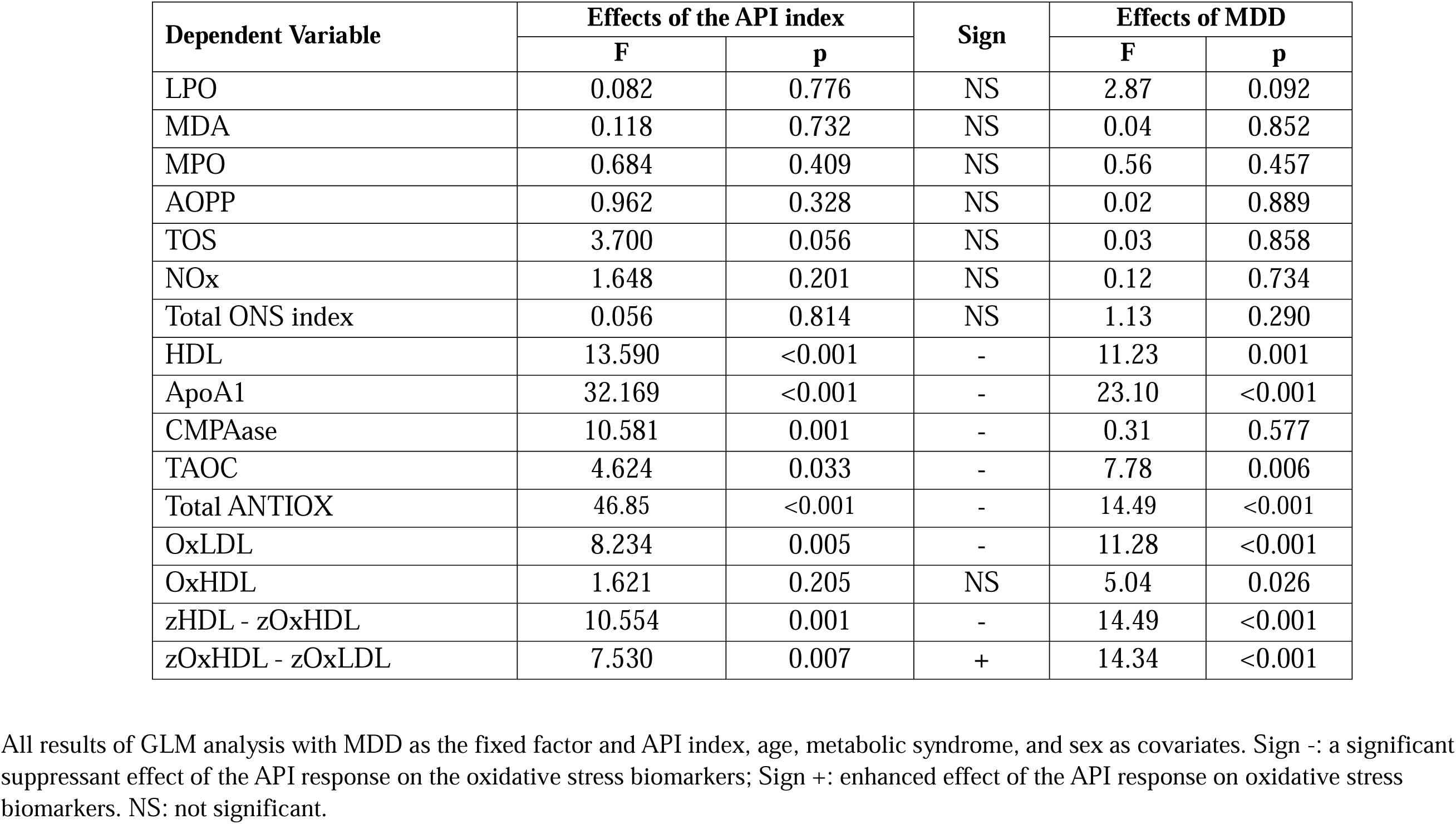
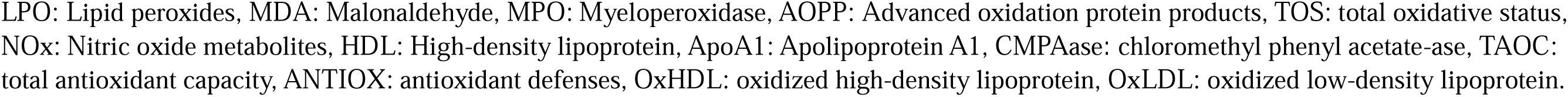
Effect of the acute phase inflammatory (API) index on oxidative stress biomarkers in major depression (MDD)

| Dependent Variable | Effects of the API index |  | Sign | Effects of MDD |  |
| --- | --- | --- | --- | --- | --- |
|  | F | p |  | F | p |
| LPO | 0.082 | 0.776 | NS | 2.87 | 0.092 |
| MDA | 0.118 | 0.732 | NS | 0.04 | 0.852 |
| MPO | 0.684 | 0.409 | NS | 0.56 | 0.457 |
| AOPP | 0.962 | 0.328 | NS | 0.02 | 0.889 |
| TOS | 3.700 | 0.056 | NS | 0.03 | 0.858 |
| NOx | 1.648 | 0.201 | NS | 0.12 | 0.734 |
| Total ONS index | 0.056 | 0.814 | NS | 1.13 | 0.290 |
| HDL | 13.590 | <0.001 | - | 11.23 | 0.001 |
| ApoA1 | 32.169 | <0.001 | - | 23.10 | <0.001 |
| CMPAase | 10.581 | 0.001 | - | 0.31 | 0.577 |
| TAOC | 4.624 | 0.033 | - | 7.78 | 0.006 |
| Total ANTIOX | 46.85 | <0.001 | - | 14.49 | <0.001 |
| OxLDL | 8.234 | 0.005 | - | 11.28 | <0.001 |
| OxHDL | 1.621 | 0.205 | NS | 5.04 | 0.026 |
| zHDL - zOxHDL | 10.554 | 0.001 | - | 14.49 | <0.001 |
| zOxHDL - zOxLDL | 7.530 | 0.007 | + | 14.34 | <0.001 |

All results of GLM analysis with MDD as the fixed factor and API index, age, metabolic syndrome, and sex as covariates. Sign -: a significant suppressant effect of the API response on the oxidative stress biomarkers; Sign +: enhanced effect of the API response on oxidative stress biomarkers. NS: not significant.
LPO: Lipid peroxides, MDA: Malonaldehyde, MPO: Myeloperoxidase, AOPP: Advanced oxidation protein products, TOS: total oxidative status, NOx: Nitric oxide metabolites, HDL: High-density lipoprotein, ApoA1: Apolipoprotein A1, CMAAse: chloromethyl phenyl acetate-ase, TAOC: total antioxidant capacity, ANTIOX: antioxidant defenses, OxHDL: oxidized high-density lipoprotein, OxLDL: oxidized low-density lipoprotein.

### Multiple regression analyses

**Table 6** summarizes the multiple regression models predicting various MDD features. In Model #1, 31.9% of the variance in OSOD was explained by zOxHDL–zOxLDL (positively), Total ANTIOX and TAOC (both negatively). After allowing for the effects of API and ACEs, model #2 demonstrated that 50.2% of the variance in OSOD was predicted by the sum of four ACEs, zOxHDL – zOxLDL ratio, and the API index (all positively), with TAOC and ApoA1 contributing negatively. Without API and ACEs, model #3 showed that 29.5% of the variance in physiosomatic symptoms was predicted negatively by Total ANTIOX, TAOC, and AOPP, and positively by zOxHDL – zOxLDL. A second model for physiosomatic symptoms (Model #4, after allowing for the effects of API and ACEs) indicated that 42.7% of its variance was explained by the sum of four ACEs, sexual abuse, and the zOxHDL – zOxLDL ratio (positively), and by TAOC and ApoA1 (negatively). In the restricted MDD group (model #5), we found that 22.5% of the variance in ROI was predicted positively by the sum of four ACEs and negatively by the Total ANTIOX index. In the restricted MDD group, ROI was also significantly and inversely associated with CMPAase activity (R² = 0.088, F = 10.68, df = 1/110, p = 0.001).

**Table 6.**
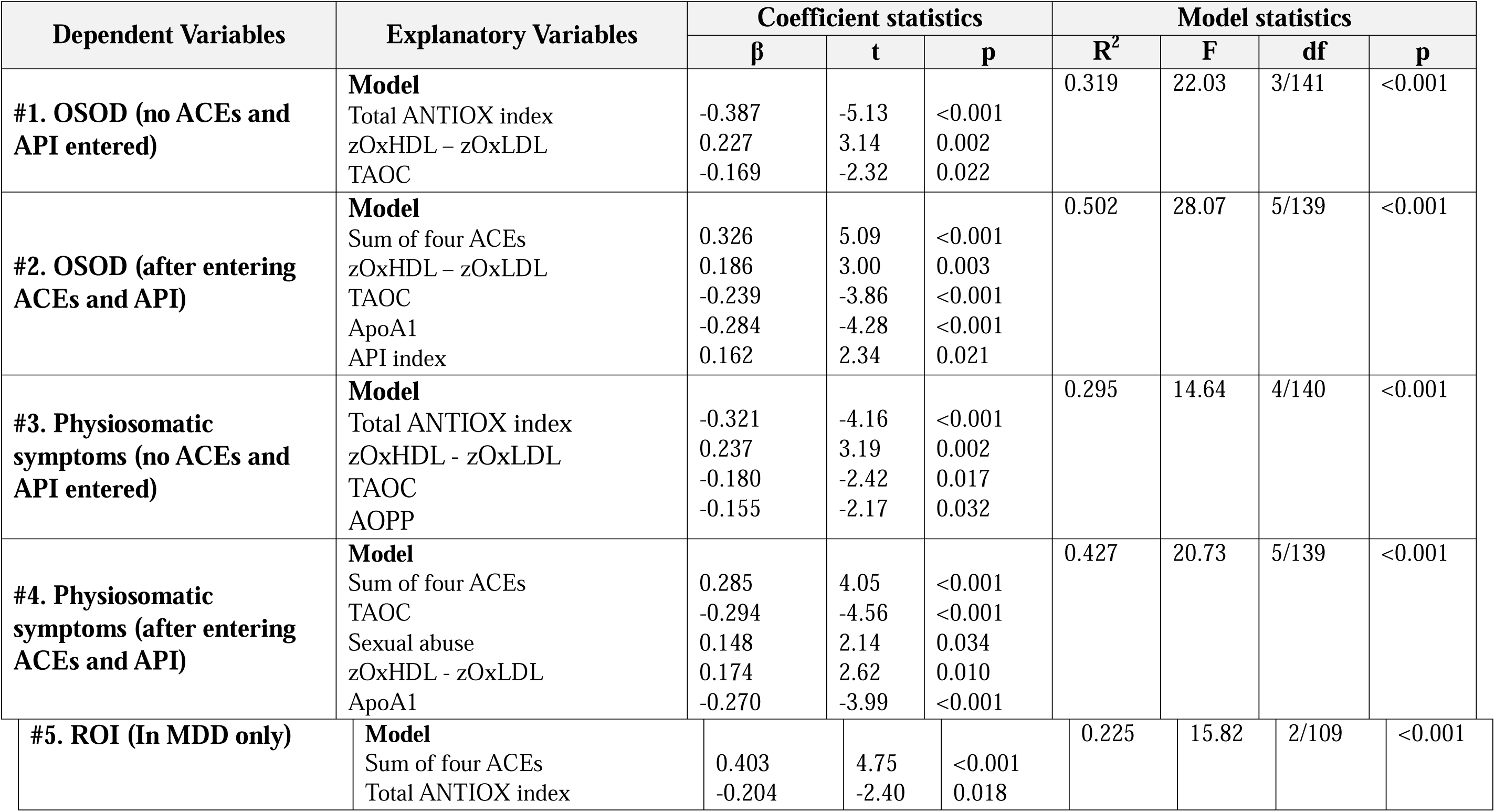
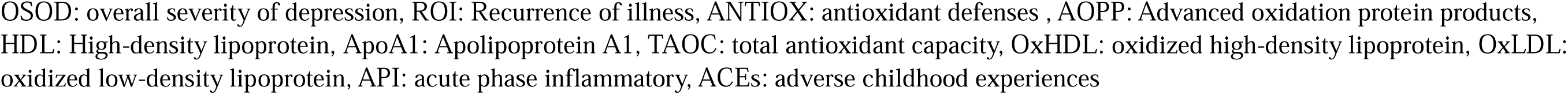
Results of multiple regression analysis with severity of depression score as dependent variables and oxidative stress biomarkers and clinical data as explanatory variables.

| Dependent Variables | Explanatory Variables | Coefficient statistics |  |  | Model statistics |  |  |  |
| --- | --- | --- | --- | --- | --- | --- | --- | --- |
| | | $\beta$ | t | p | R<sup>2</sup> | F | df | p |
| #1. OSOD (no ACEs and API entered) | **Model** |  |  |  | 0.319 | 22.03 | 3/141 | <0.001 |
|  | Total ANTIOX index | -0.387 | -5.13 | <0.001 |  |  |  |  |
|  | zOxHDL – zOxLDL | 0.227 | 3.14 | 0.002 |  |  |  |  |
|  | TAOC | -0.169 | -2.32 | 0.022 |  |  |  |  |
| #2. OSOD (after entering ACEs and API) | **Model** |  |  |  | 0.502 | 28.07 | 5/139 | <0.001 |
|  | Sum of four ACEs | 0.326 | 5.09 | <0.001 |  |  |  |  |
|  | zOxHDL – zOxLDL | 0.186 | 3.00 | 0.003 |  |  |  |  |
|  | TAOC | -0.239 | -3.86 | <0.001 |  |  |  |  |
|  | ApoA1 | -0.284 | -4.28 | <0.001 |  |  |  |  |
|  | API index | 0.162 | 2.34 | 0.021 |  |  |  |  |
| #3. Physiosomatic symptoms (no ACEs and API entered) | **Model** |  |  |  | 0.295 | 14.64 | 4/140 | <0.001 |
|  | Total ANTIOX index | -0.321 | -4.16 | <0.001 |  |  |  |  |
|  | zOxHDL - zOxLDL | 0.237 | 3.19 | 0.002 |  |  |  |  |
|  | TAOC | -0.180 | -2.42 | 0.017 |  |  |  |  |
|  | AOPP | -0.155 | -2.17 | 0.032 |  |  |  |  |
| #4. Physiosomatic symptoms (after entering ACEs and API) | **Model** |  |  |  | 0.427 | 20.73 | 5/139 | <0.001 |
|  | Sum of four ACEs | 0.285 | 4.05 | <0.001 |  |  |  |  |
|  | TAOC | -0.294 | -4.56 | <0.001 |  |  |  |  |
|  | Sexual abuse | 0.148 | 2.14 | 0.034 |  |  |  |  |
|  | zOxHDL - zOxLDL | 0.174 | 2.62 | 0.010 |  |  |  |  |
|  | ApoA1 | -0.270 | -3.99 | <0.001 |  |  |  |  |
| #5. ROI (In MDD only) | **Model** |  |  |  | 0.225 | 15.82 | 2/109 | <0.001 |
|  | Sum of four ACEs | 0.403 | 4.75 | <0.001 |  |  |  |  |
|  | Total ANTIOX index | -0.204 | -2.40 | 0.018 |  |  |  |  |

OSOD: overall severity of depression, ROI: Recurrence of illness, ANTIOX: antioxidant defenses , AOPP: Advanced oxidation protein products, HDL: High-density lipoprotein, ApoA1: Apolipoprotein A1, TAOC: total antioxidant capacity, OxHDL: oxidized high-density lipoprotein, OxLDL: oxidized low-density lipoprotein, API: acute phase inflammatory, ACEs: adverse childhood experiences

## Discussion

### Reduced HDL levels and Elevated OxHDL in MDD

The first major finding of this study is that MDD is characterized by decreased HDL levels coupled with elevated OxHDL in patients relative to healthy controls. These findings extend those of earlier research indicating reduced HDL levels in MDD (Maes, Smith et al. 1997, Almulla, Thipakorn et al. 2023, Jirakran, Almulla et al. 2025) and present oxidative modification of HDL as a novel characteristic of the condition. In individuals with MetS, HDL levels were decreased; however, OxHDL levels did not differ between the MetS and non-MetS groups. This suggests that HDL particle oxidation in MDD is not influenced by MetS comorbidity.

The increase in OxHDL may explain the decrease in measurable HDL, as oxidative damage affects HDL stability and hastens its functional deterioration. HDL oxidation occurs via the formation of HOCl derived from MPO and peroxidation mediated by 15-LOX, both of which alter ApoA-I and hinder ABCA1-dependent cholesterol efflux (Bergt, Pennathur et al. 2004, Matsunaga, Hara et al. 2010, Shao, Oda et al. 2010). The observed increase in OxHDL in this study occurred independently of changes in MPO and LPO, indicating a specific susceptibility of HDL to oxidative remodeling, rather than a broad occurrence of lipid peroxidation. This selective oxidation aligns with the established effects of H O -induced modifications of HDL, which negatively affect ApoA1 and decrease PON1 activity (Fadaei and Davies 2022, Napolitano, Fasciolo et al. 2023). Non-enzymatic processes such as glycation, carbamylation, and proteolytic degradation contribute to the deterioration of HDL structure (Ferretti, Bacchetti et al. 2006, Santana and Brown 2018).

The implications of elevated OxHDL might be extensive and significant in clinical contexts. OxHDL exhibits a reduction in antioxidant and anti-inflammatory capabilities, promotes the production of reactive oxygen species (ROS), elevates cytokine release, activates matrix metalloproteinases, and hinders endothelial repair (Soumyarani and Jayakumari 2012, Wu, He et al. 2015, Xepapadaki, Zvintzou et al. 2020). Oxidative modifications of ApoA1, along with decreased PON1 and LCAT activity, impair HDL maturation, restrict cholesterol efflux, and diminish NO-dependent endothelial protection (DiDonato, Huang et al. 2013, Hewing, Parathath et al. 2014, Huang, Yancey et al. 2020). OxHDL enhances oxidative injury in neurons, astrocytes, and microglia within the central nervous system, resulting in cell death (Keller, Hanni et al. 2000). The observed consequences indicate that elevated OxHDL serves not merely as a marker of oxidative stress but might also function as an active mediator of NIMETOX dysfunctions.

HDL, PON1, and ApoA1 are fundamental components of RCT that modulate NF-κB signaling, Toll-like receptor 4 (TLR-4) activity, complement pathways, and T-cell responses (Camps, Iftimie et al. 2017, Morris, Puri et al. 2021, Morris, Gevezova et al. 2022). Thus, decreased HDL and elevated OxHDL consequently impair various immunoregulatory systems pertinent to MDD. Moreover, HDL transports miRNAs that regulate lipid transporters (Ouimet and Moore 2013, Scherrer, Zago et al. 2015), and its oxidative modification could disrupt miRNA-mediated metabolic signaling. The observed convergent disturbances suggest that HDL oxidation serves as a connection between oxidative stress, immune activation, and lipid dysfunction in MDD.

### The HDL to OxHDL ratio

The zHDL - zOxHDL ratio identified in this study functions as a significant biomarker for HDL vulnerability to oxidative stress. This index reflects the extent of structural compromise in HDL particles. The significant correlation between the API index, characterized by elevated mCRP and decreased albumin and transferrin (Maes, Niu et al. 2025), and alterations in HDL and OxHDL support the hypothesis that inflammation is a contributor to HDL dysfunction in MDD (Chen, Niu et al. 2025). Collectively, our results suggest that diminished HDL and elevated OxHDL reflect a consistent pattern of inflammatory and oxidative damage. The combination of both biomarkers indicates a selective oxidation of HDL particles with impairments of its essential components, namely ApoA1, PON1 and LCAT. This positions the zHDL - zOxHDL ratio as a significant biomarker for oxidative vulnerability to HDL particles associated with MDD.

### Decreased OxLDL Levels in MDD

This study’s second major finding is the significant reduction of measurable OxLDL in patients with MDD compared to healthy controls. In contrast, previous studies have reported elevated OxLDL levels in MDD (Ogłodek 2017), while a recent meta-analysis has indicated unchanged levels (Almulla, Thipakorn et al. 2023). In Alzheimer’s disease, reduced levels of OxLDL were observed (Grossi, Carvalho et al. 2018). A decrease in measurable serum OxLDL does not signify a reduction in oxidative stress but instead might reflect a change in the biological distribution of oxidized LDL. This interpretation is corroborated by consistent evidence of elevated IgG targeting OxLDL in MDD (Maes, Simeonova et al. 2019, Almulla, Thipakorn et al. 2023), suggesting that OxLDL is sequestered into immune complexes instead of being present in its free, quantifiable form. Therefore, diminished measurable OxLDL in MDD suggests enhanced oxidative modification of LDL and increased autoimmune recognition of oxidation-specific epitopes.

Decreased circulating OxLDL is detrimental as it likely indicates increased Lectin-like oxidized low-density lipoprotein receptor-1 (LOX-1)–mediated uptake of OxLDL into endothelial cells, macrophages, and smooth muscle cells. LOX-1 expression is elevated in response to oxidative stress, pro-inflammatory cytokines, and TGF-β1, all of which are consistently elevated in MDD (Minami, Kume et al. 2000, Jovanovic, Mitkovic-Voncina et al. 2021, Gędek, Modrzejewski et al. 2025). Elevated LOX-1 activity enhances the internalization of OxLDL, resulting in a reduction of its detectable plasma levels and facilitating its accumulation in vascular tissues. This interpretation aligns with observations indicating that OxLDL accumulates in plaques at concentrations approximately 70 times greater than in plasma (Nishi, Itabe et al. 2002). Consequently, reduced levels of circulating OxLDL do not confer protection; instead, they may indicate heightened vascular oxidative damage and the onset of atherogenic remodeling in MDD. This pattern yields detrimental consequences. Internalized OxLDL leads to endothelial dysfunction, facilitates macrophage foam cell formation, enhances inflammatory gene expression, and activates NF-κB and JNK signaling (Sun and Chen 2011). The interaction between LOX-1 and PCSK9 enhances the uptake of OxLDL and promotes inflammation (Ding, Liu et al. 2015).

Moreover, HDL is highly susceptible to oxidative modification, often more so than LDL, due to its lower antioxidant content and the presence of oxidation-sensitive residues in ApoA-I and ApoA-II (Garner, Witting et al. 1998). The altered zOxHDL - zOxLDL ratio observed in this study reflects a clear imbalance in lipid oxidation processes in MDD, integrating the increase in OxHDL discussed earlier with the reduction in measurable OxLDL. The pattern suggests a shift toward greater oxidative modification of HDL relative to LDL, making the zOxHDL - zOxLDL ratio a meaningful biomarker of altered lipid-oxidative dynamics in MDD.

In this study, the API index demonstrated a significant inverse correlation with reduced OxLDL levels and an increase in the zOxHDL - zOxLDL ratio. This pattern suggests that inflammation directly contributes to the reduction of measurable OxLDL, likely through the promotion of immune complex formation and the enhancement of LOX-1–mediated clearance. Concurrently, inflammation increases oxidative damage to HDL particles, thereby altering the zOxHDL - zOxLDL balance towards a state of increased oxidative susceptibility.

Our results indicated no significant difference in OxLDL levels or the zOxHDL - zOxLDL ratio between participants with and without MetS, suggesting that the reduction of OxLDL in MDD is independent of metabolic status. This finding distinguishes OxLDL biology from the MetS-associated abnormalities commonly associated with MDD. The findings support the conclusion that the reduced measurable OxLDL is influenced by depression-related oxidative-immune mechanisms rather than lipid changes driven by MetS.

Overall, integration of reduced OxLDL, increased OxHDL, and a high zOxHDL - zOxLDL ratio reveals a characteristic oxidative lipid signature in MDD, indicating intensified inflammatory activity, enhanced vascular uptake of oxidized LDL, and increased cardiovascular vulnerability.

### Decreased Antioxidant Levels and Unchanged Lipid Peroxidation Markers

The present study’s third key finding shows that MDD is characterized by markedly reduced antioxidant defenses, including lower ApoA1, CMPAase, TAOC, and the Total ANTIOX composite score, while classical lipid peroxidation and protein oxidation markers such as LPO, MDA, MPO, TOS, AOPP, NOx, and the total ONS score remained unchanged. Reduced ApoA1, diminished CMPAase, and lower PON1 and LCAT activities have been documented in MDD and align closely with the present findings (Maes, Jirakran et al. 2025). Furthermore, meta-analytic evidence indicates widespread reductions in RCT-related antioxidant functions, vitamin-associated antioxidant defenses, ApoE, and ghrelin (Almulla, Thipakorn et al. 2023), as well as decreased vitamin E, CoQ10, zinc, glutathione peroxidase, and overall antioxidant status in MDD (Maes, Galecki et al. 2011). Previous work has shown stable LPO levels but elevations in specific oxidative products such as MDA, 8-isoprostanes, 4-HNE, and peroxides (Almulla, Thipakorn et al. 2023), while milder depression may even show reduced lipid hydroperoxides levels (Brinholi, Vasupanrajit et al. 2024).

The present study also found that MetS selectively reduced ApoA1 but did not affect CMPAase or the total ANTIOX composite score. This suggests that the core antioxidant deficits identified here are primarily depression-related rather than driven by metabolic abnormalities. ApoA1, TAOC, and the total ANTIOX composite remained significantly lowered even after adjusting for inflammation, indicating that these antioxidant deficits persist independently of the acute phase response.

Overall, lowered antioxidant levels appear to be a key factor in MDD and the consequent increase in oxidative stress might target the most vulnerable HDL particles rather than LDL particles or other lipids and proteins. Damaged HDL particles are dysfunctional further impairing the antioxidant defenses and the RCT pathway, thereby causing activated neuro-immune pathways and other NIMETOX pathways (Maes, Almulla et al. 2025).

### Prediction of MDD and its clinical phenome

The fourth major finding of this research demonstrates that the integration of HDL-related biomarkers with the API index achieved cross-validated diagnostic accuracies ranging from 79% to 83%. This indicates that up to 83% of MDD patients display antioxidant and inflammatory aberrations with a specificity of around 83%. This pattern is consistent with recent multimarker studies. Almulla, Niu et al. (2025) reported an AUC of 0.855 utilizing mCRP, albumin, transferrin, IL-17A, and ACE domains (Almulla, Niu et al. 2025). Chen, Niu et al. (2025) demonstrated that the integration of the API index with total cholesterol, ApoA1, and BMI achieved an accuracy of 81.7% in distinguishing MDD (Chen, Niu et al. 2025). Excluding subjects with MetS in these studies somewhat enhanced specificity, suggesting that MetS status may affect the accuracy of the model. The current findings align with this trend, indicating that comprehensive integration of inflammatory, oxidative, and lipid markers yields the most consistent discriminatory performance (Maes, Almulla et al. 2025).

Our findings also indicate that the clinical phenome of MDD—including ROI, OSOD and physiosomatic symptoms—is to a large extent explained by elevated OxHDL, decreased OxLDL, and lower levels of antioxidants. Previous research indicates that OSOD and physiosomatic symptoms are related to ACEs, negative APPs, and cytokine levels (Niu, Zhang et al. 2025), ACEs, ApoA1, transferrin, mCRP, and HOMA-IR (Almulla, Niu et al. 2025, Chen, Niu et al. 2025, Luo, Niu et al. 2025). These associations indicate that NIMETOX disturbances directly contribute to the manifestation of various clinical domains in MDD.

The current study showed that increasing ROI is accompanied by lowered antioxidant defenses and lower CMPAase activity, which protects HDL particles against oxidative damage (Moreira, Boll et al. 2019). Previously, it was shown that lowered PON1 activity is associated with increased number of depressive and manic episodes (Moreira, Correia et al. 2019). The findings correspond with evidence from Maes et al. (Maes 2022, Maes, Rachayon et al. 2022, Maes and Stoyanov 2022, Maes, Moraes et al. 2023, Maes, Rachayon et al. 2023, Maes, Vasupanrajit et al. 2023, Maes, Zhou et al. 2024, Maes, Zhou et al. 2024), showing that the progression of ROI reflects enhanced activation of NIMETOX pathways, which include lowered antioxidant defenses, immune shifts, lipid disturbances, and oxidative injuries.

Finally, we observed that ACEs (neglect and abuse but not sexual abuse) impacted not only the MDD phenome (OSOD, physiosomatic symptoms and ROI) but also antioxidant values, including ApoA1 and CMPAase. Previously, some papers showed that ACEs impact antioxidant levels and may increase lipid peroxidation levels in subjects with MDD (Maes, Moraes et al. 2021, Brinholi, Vasupanrajit et al. 2024). The latter study showed that ACEs are associated with antioxidants and oxidative stress in the first episode of MDD (Brinholi, Vasupanrajit et al. 2024), whereas this condition is not accompanied by immune activation (Maes, Vasupanrajit et al. 2025). This may suggest that aberrations in ACE-associated aberrations in redox mechanisms, rather than immune processes, are associated with the onset of MDD.

## Limitations

The current findings have several limitations that require consideration. The case–control design limits the ability to make strict causal inferences about the relationship between lipoprotein oxidation and the progression of MDD. This study quantified OxHDL and OxLDL for the first time in this context; however, it did not include autoantibodies such as IgG and IgM that target these neoepitopes (Almulla, Thipakorn et al. 2023). Documented differences in oxidative stress profiles between Chinese and Western populations, along with evidence that latitude influences RCT, HDL, and lipid-associated antioxidants (Almulla, Thipakorn et al. 2023), highlight the need for replication in other regions. Finally, the integrated biomarker models exhibited strong discriminatory performance. However, validation in more diverse samples is necessary to confirm their generalizability across different clinical and demographic contexts.

## Conclusions

Our study provides the first evidence that MDD is accompanied by increased OxHDL, elevated zHDL - zOxHDL and zOxHDL – zOxLDL ratios, and reductions in both HDL and OxLDL, independent of MetS and the API response. We also identified diminished antioxidant defenses despite unchanged classical lipid peroxidation markers. Most importantly, we delineated a selective pattern of HDL particle oxidation rather than a generalized rise in oxidative damage markers or damage to LDL or other lipids. In addition, the integration of HDL particle markers together with API markers shows strong diagnostic and predictive potential for MDD. These data suggest that OxHDL and OxLDL-related indices represent new components within the NIMETOX framework and may serve as promising targets for future therapeutic development.

## Supporting information

Electronic Supplementary File

## Data Availability

The corresponding author Michael Maes will grant access to the dataset supporting this study upon reasonable request, following a thorough data review.

## Acknowledgements

Not Applicable

## Ethical approval and consent to participate

The Research Ethics Committee at Sichuan Provincial People’s Hospital in Chengdu, China, approved the study (Ethics Approval No. 2024-203). All participants provided written informed consent prior to enrollment.

## Author’s contributions

All authors contributed equally to the research and approved the final manuscript.

## Availability of data

The corresponding author (MM) will grant access to the dataset supporting this study upon reasonable request, following a thorough data review.

