## Supplementary material for "Increased Oxidation Susceptibility of HDL Particles as a Mechanistic Signature of Major Depressive Disorder": Electronic Supplementary File

**ELECTRONIC SUPPLEMENTARY FILE (ESF)**

**ESF, Table 1.** Methods used to assay the biomarkers in the present study

|  | **Assay** | **Method and kit** | **Equipment** |
| --- | --- | --- | --- |
| 1 | Albumin | Immunoturbidimetric assay (DIAYS DIAGNOSTIC SYSTEM (SHANGHAI) CO., LTD). Sensitivity 0.03 g/L, intra-assay and inter-assay analytical coefficients of variation (CVs) of 1.96% and 0.67%, respectively. | Fully automated biochemical analyzer (ADVIA 2400, Siemens Healthcare Diagnostic Inc) |
| 2 | Transferrin | Bromocresol green method (Beijing Strong Biotechnologies, Inc.). The sensitivity is 0.03 g/L, with the intra-assay and inter-assay analytical CVs of 1.20% and 2.10%, respectively. | Fully automated biochemical analyzer (ADVIA 2400, Siemens Healthcare Diagnostic Inc) |
| 3 | Low-density lipoprotein cholesterol (LDL) | Direct method-surfactant clearance method (Beijing Strong Biotechnologies, Inc.) with the intra-assay and inter-assay CVs of < 3% and < 10%, respectively. | Fully automated biochemical analyzer (ADVIA 2400, Siemens Healthcare Diagnostic Inc) |
| 4 | High-density lipoprotein cholesterol (HDL) | Direct method-select inhibition method (Beijing Strong Biotechnologies, Inc.) with the intra-assay and inter-assay CVs of < 4% and < 10%, respectively. | Fully automated biochemical analyzer (ADVIA 2400, Siemens Healthcare Diagnostic Inc) |
| 5 | Apolipoprotein A1 (ApoA1) | Immunoturbidimetric method (Beijing Strong Biotechnologies, Inc.) with the intra-assay and inter-assay CVs of < 3% and < 10%, respectively. | Fully automated biochemical analyzer (ADVIA 2400, Siemens Healthcare Diagnostic Inc) |
| 9 | Monomeric C-reactive protein (mCRP) | ELISA (BioVendor, Brno, Czech Republic). The sensitivity is 0.63 ng/mL, and the intra-assay and inter-assay CV is < 10% and < 15%, respectively. | Microplate reader (Thermofisher, Sky-High) |
| 10 | Paraoxonase (PON)1 (Chloromethyl)phenyl acetate CMPAase | Serum CAMPAse activity was quantified by monitoring the hydrolysis of 4- (CMPA, CAS No.: 39720-27-9, Merck, USA) at 280 nm(Brinholi et al. 2024; Maes et al. 2022). The reactions were conducted in UV-transparent 96-well microplates at 25℃, with kinetic absorbance recorded for 4 minutes (16 measurements at 15-second intervals). CMPAse activity was calculated in U/mL based on the molar extinction coefficients of 1.30 mmol/L·cm-1. | Microplate reader (Thermofisher, Sky-High) |
| 11 | Lipid peroxide (LPO) | LPO was measured using colorimetric assay kits (Elabscience, E-BC-K176-M, Wuhan, China). The sensitivity is 0.70 μmol/L. The intra-assay and inter-assay CVs are 3.1% and 3.5%, respectively. | Microplate reader (Thermofisher, Sky-High) |
| 12 | Nitric oxide metabolites (NOx) | Nox was measured by the nitrate reductase method (Elabscience, E-BC-K135-M, Wuhan, China). The sensitivity is 1.38 μmol/L. The intra-assay and inter-assay CVs are 6% and 8%, respectively. | Microplate reader (Thermofisher, Sky-High) |
| 13 | Myeloperoxidase (MPO) | MPO activity was determined by microassay (Solarbio, BC5715, Beijing, China). The sensitivity is 1.13 μmol/L, and the intra-assay and inter-assay CVs are 4.1% and 7.2%, respectively. | Microplate reader (Thermofisher, Sky-High) |
| 14 | Malondialdehyde (MDA) | MDA was measured using colorimetric assay kits (TBA method) (Elabscience, E-BC-K025-M, Wuhan, China). | Microplate reader (Thermofisher, Sky-High) |
| 15 | Advanced oxidation protein products (AOPP) | ELISA (Ruixin Biotech, RX105143H, Wuhan, China). The sensitivity is 12.5 ng/mL. The intra-assay and inter-assay CVs are 5% and 8%, respectively. | Microplate reader (Thermofisher, Sky-High) |
| 16 | Total oxidative status (TOS) | TOS was measured using colorimetric assay kits (Elabscience, E-BC-K802-M, Wuhan, China). The sensitivity is 2.5 μmol H_2_O_2_ Equiv./L. The intra-assay and inter-assay CVs are 2.3% and 3.5%, respectively. | Microplate reader (Thermofisher, Sky-High) |
| 17 | Total antioxidant capacity (TAOC) | TAOC was measured using colorimetric assay kits (Elabscience, E-BC-K136-M, Wuhan, China). The sensitivity is 0.62 U/mL. The intra-assay and inter-assay CVs are 4.8% and 5.6%, respectively. | Microplate reader (Thermofisher, Sky-High) |
| 18 | Oxidized high-lipoprotein (Ox-HDL) | ELISA (Fine Test, EH4858, Wuhan, China). The sensitivity is 0.938 ng/mL, and the intra-assay and inter-assay CVs of 5.12% and 5.22%, respectively. | Microplate reader (Thermofisher, Sky-High) |
| 19 | Oxidized low-lipoprotein (Ox-LDL) | ELISA (Elabscience, E-EL-H6021, Wuhan, China). The sensitivity is 37.5 pg/mL, and the intra-assay and inter-assay CVs of 6.62% and 7.04%, respectively. | Microplate reader (Thermofisher, Sky-High) |

**ESF, Table 2**. Socio-demographic, clinical and biomarker data of patients with major depression (MDD) and healthy controls (HC).

| **Variables** | **HC (n = 40)** | **MDD (n = 125)** | **F / χ²** | **df** | **p-value** |
| --- | --- | --- | --- | --- | --- |
| Age (years) | 37.1 (13.7) | 35.7 (12.1) | 0.37 | 1/163 | 0.542 |
| Gender (m/f) | 13/27 | 38/87 | 0.06 | 1 | 0.845 |
| History of smoking (No/Yes) | 37/3 | 101/24 | 3.03 | 1 | 0.091 |
| Metabolic syndrome (MetS) (No/Yes) | 29/10 | 102/22 | 1.17 | 1 | 0.355 |
| Ranking MetS | 1.590 (1.464) | 1.419 (1.257) | 0.50 | 1/161 | 0.479 |
| Body Mass Index (BMI) (kg/m²) | 23.5 (4.1) | 22.3 (3.4) | 3.58 | 1/163 | 0.060 |
| Waist circumference (WC) (cm) | 79.4 (11.7) | 78.3 (11.3) | 0.28 | 1/163 | 0.595 |
| zBMI + zWC (z score) | 0.332 (2.020) | -0.106 (1.840) | 1.64 | 1/163 | 0.202 |
| Overall severity of depression (z score) | -1.465 (0.260) | 0.537 (0.510) | MWUT | - | <0.001 |
| Physiosomatic symptoms ( z scores) | -1.300 (0.421) | 0.477 (0.671) | MWUT | - | <0.001 |
| Recurrence of illness index (z score) | -2.559 (0.356) | 0.819 (2.140) | MWUT | - | <0.001 |
| Sum Four ACEs | 30.98 (9.19) | 41.15 (11.78) | 24.93 | 1/163 | <0.001 |
| Sexual abuse | 5.23 (0.58) | 6.34 (2.46) | 8.08 | 1/163 | 0.005 |

All results are shown as mean (SD). Results of analysis of variance and analysis of contingency tables. MWUT: Mann-Whitney U test.

**ESF, Table 3.** The biomarkers measured in the current study in people with and without metabolic syndrome (MetS)

| **Variables (z scores)** | **No MetS (n=131)** | **MetS (n=31)** | **F value**  **(df=1/145)** | **p-value** |
| --- | --- | --- | --- | --- |
| Albumin | -0.001 (0.083) | -0.031 (0.188) | 0.02 | 0.890 |
| Transferrin | 0.008 (0.084) | 0.032 (0.191) | 0.01 | 0.912 |
| Monomeric C-reactive protein | -0.026 (0.090) | 0.137 (0.365) | 0.49 | 0.485 |
| Acute phase inflammatory (API) index | -0.033 (0.162) | 0.136 (0.365) | 0.16 | 0.686 |
| LPO | 0.039 (0.109) | 0.446 (0.190) | 3.39 | 0.067 |
| MDA | -0.105 (0.113) | 0.157 (0.197) | 1.30 | 0.256 |
| MPO | 0.020 (0.115) | 0.187 (0.201) | 0.51 | 0.476 |
| AOPP | 0.136 (0.111) | 0.122 (0.193) | 0.00 | 0.952 |
| TOS | 0.031 (0.104) | 0.470 (0.181) | 4.38 | 0.038 |
| Nitric oxide metabolites | 0.023 (0.112) | 0.018 (0.196) | 0.00 | 0.983 |
| Total ONS index | 0.047 (0.107) | 0.551 (0.186) | 5.43 | 0.021 |
| HDL | 0.308 (0.094) | -0.668 (0.164) | 26.10 | <0.001 |
| ApoA1 | 0.339 (0.093) | -0.292 (0.163) | 11.13 | 0.001 |
| CMPAase | 0.047 (0.110) | 0.389 (0.192) | 2.36 | 0.126 |
| TAOC | 0.126 (0.100) | 0.680 (0.174) | 7.46 | 0.007 |
| Total ANTIOX index | 0.350 (0.099) | 0.030 (0.172) | 2.57 | 0.111 |
| zTotal OS – zTotal ANTIOX | -0.225 (0.102) | 0.368 (0.178) | 8.22 | 0.005 |
| OxHDL | -0.033 (0.090) | 0.135 (0.203) | 0.52 | 0.470 |
| zHDL - zOxHDL (Zscore) | 0.298 (0.100) | -0.490 (0.175) | 15.07 | <0.001 |
| OxLDL | 0.028 (0.086) | -0.108 (0.195) | 0.37 | 0.542 |
| zOxHDL - zOxLDL (Zscore) | -0.204 (0.107) | -0.184 (0.186) | 0.01 | 0.927 |

All data are shown as marginal estimated mean (SE); results of GLM analysis with age, sex, body mass index and major depression as covariates. HDL, ApoA1, TAOC, z Total OS – z TotalANTIOX, and zHDL – zOxHDL remained significantly different between both groups after False Discovery Rate p correction,

LPO: Lipid peroxides, MDA: Malonaldehyde, MPO: Myeloperoxidase, AOPP: Advanced oxidation protein products, TOS: total oxidative status, CMPAase: chloromethyl phenyl acetate, TAOC: total antioxidant capacity, ONS: oxidative and nitrosative stress; ANTIOX: antioxidant defenses, oxHDL: oxidized high-density lipoprotein, oxLDL: oxidized low-density lipoprotein.
